# External validation of a risk score model for predicting major clinical events in adults after atrial switch operation for transposition of the great arteries

**DOI:** 10.1101/2023.03.31.23286966

**Authors:** Mathieu Albertini, Beatrice Santens, Flavia Fusco, Berardo Sarubbi, Pastora Gallego, Maria-Jose Rodriguez-Puras, Katja Prokselj, Martijn Kauling, Jolien Roos-Hesselink, Fabien Labombarda, Alexander Van De Bruaene, Werner Budts, Laurence Iserin, Odilia Woudstra, Berto Bouma, Magalie Ladouceur

## Abstract

**Background:** A risk model has been proposed to provide a patient individualized estimation of risk for major clinical events (heart failure, ventricular arrhythmia, all-cause mortality) in patients with transposition of the great arteries (D-TGA) repaired by an atrial switch operation. The aim of this study was to externally validate the model.

**Methods:** A retrospective, multicentric, longitudinal cohort of 417 patients with D-TGA (median age 24 years at baseline [interquartile range 18-30], 63% male) independent of the model development and internal validation cohort was studied. Data on risk model predictors (age >30 years, prior ventricular arrhythmia, age >1 year at atrial switch, moderate or severe right ventricular dysfunction, severe tricuspid regurgitation and at least mild left ventricular (LV) dysfunction) were collected from the time of baseline clinical evaluation. The performance of the prediction model in predicting risk at 5 years was assessed.

**Results:** Twenty-five patients (5.9%) met the major clinical events endpoint within 5 years. Model validation showed a good discrimination between high and low 5-year risk patients (Harrell’s C-index of 0.73 (95% CI 0.65–0.81)) but tended to overestimate this risk (calibration slope of 0.20 (95% CI 0.03–0.36)). We separately evaluated predictors of major clinical events in our cohort. History of heart failure and at least mildly impaired sub pulmonary LV function remained the strongest predictors of major clinical events in this population.

**Conclusions:** We reported the first external validation of a major clinical events risk model in a large D-TGA patient population. Although a good discrimination, the model tends to overestimate the absolute 5-year risk. Subpulmonary LV dysfunction appears to be a key marker in the prognosis of patients with Senning and Mustard. Further optimizing risk models are needed to individualize risk predictions in D-TGA patients.

**Clinical Perspective:** **What is new?**

This is the first external validation of a risk model for major clinical events in D-TGA patients after atrial switch and the largest study emphasizing the importance of assessing subpulmonary left ventricle (LV) function in these patients.

**What are clinical implications?**

Risk model for major clinical events can be used to discriminate patients at low from those at intermediate and high risk. This tool helps determine follow-up intensity, and support management decisions specifically for intermediate- and high-risk patients with a history of heart failure and at least mild sub pulmonary left ventricle (LV) dysfunction.

Sub-pulmonary LV, which can be the “forgotten chamber” in these patients with a systemic right ventricle, should be carefully and regularly surveyed.

Patients from the intermediate/high-risk group with history of heart failure, and subpulmonary LV dysfunction have a poor prognosis and should be referred for consideration of advanced therapies

## Introduction

The survival of patients with transposition of the great arteries (D-TGA) has improved dramatically after introduction of the atrial switch (AtrS) procedures by Ake Senning and William Mustard from the mid-1960s to the mid-1980s, when the use of the atrial switch procedure became commonplace. Atrial switch procedures involve redirection of the blood flow in the atria, consequently the morphological right ventricle supports the systemic circulation, whereas the left ventricle (LV) supports the pulmonary circulation. Although these procedures led to a significant improvement in survival during the first two decades of life^1^, late complications are frequently encountered including systemic right ventricular (sRV) dysfunction, arrhythmias, heart failure (HF) and sudden cardiac death^2 3 4^. The survival at 40 years is estimated to be between 70 to 80%^5 6 2^. Nowadays most of patients are approaching middle age.

Assessing the prognosis of these patients has become essential to identify patients at risk for major clinical events (HF, ventricular arrhythmia, death). This approach would allow a tailored risk prediction to support decisions for follow-up interval and therapeutic management. Patients in the category of high risk would likely benefit from referral to a quaternary center, where issues of cardiac support and heart transplantation would be discussed, considering the hardly proven effectiveness of the drug treatments in congenital heart disease (CHD) with a sRV ^7 8 9^.

Recently, Woudstra et al provided a clinical risk model that estimates the risks of major events during the clinical course of patients with D-TGA and atrial switch. The model gave a practical risk score based on 6 criteria (age >30 years, repair at >1 year, prior ventricular arrhythmia, moderate or severe RV dysfunction, severe tricuspid regurgitation and at least mild left ventricular dysfunction) stratifying patients into low, intermediate, and high-risk groups of HF, ventricular tachycardia, and death at 5 and 10 years^10^. Although this prediction model showed a good discriminatory ability on internal validation analysis, an external validation in an independent population has not yet been performed. The aim of the current study was to validate this predictive score in a large, independent, multi-center patient population.

## Methods

### Patients

The study cohort comprised adult patients (>16 years old) with a D-TGA and atrial switch from 7 European participating centres in the MARES registry (NCT03833843) ^4^ (Hôpital Européen Georges Pompidou, Paris - Erasmus Medical Center, Rotterdam – University Hospital Leuven, Leuven - Monaldi Hospital, Naples - Hospital Virgen del Rocio, Seville - University Medical Center Ljubljana, Ljubljana - Centre Hospitalo-Universitaire de Caen, Caen). The study complies with the Declaration of Helsinki and ethical or research committee approval was obtained in each contributing centre.

Included patients were evaluated between January 2000 and December 2018. We excluded patients with missing data for calculation of the major clinical event score and those with less than 3 years of follow-up. This cohort was independent of the major clinical event score development cohort ^10^.

Patients were followed up from the first hospital visit until December 2019 or the date of primary outcome. The patients’ medical records were reviewed to collect demographic information, medical and surgical details.

The potential risk factors of clinical events in TGA after AtrS corresponded to those retained in the risk prediction score developed by Woudstra et al^10^ and are listed in Table 1. Moreover, additional predictors selected on the basis of a literature review were assessed (in supplemental data Table S1^2,4,6,10–26^). Complex TGA was defined as TGA associated with ventricular septal defect (VSD), left ventricular outflow tract (LVOT) obstruction and/or aortic coarctation. Associated pulmonary arterial hypertension was noted in the presence of Eisenmenger syndrome or when pre-capillary pulmonary hypertension was invasively confirmed according to the ESC guidelines^27^. Atrial arrhythmia history encompassed all the types of supraventricular arrhythmia including ectopic atrial tachycardia, atrioventricular nodal reentrant tachycardia, atrioventricular reciprocating tachycardia, intra-atrial re-entry tachycardia, atrial flutter, and atrial fibrillation. Holter and pacemaker/ICD monitoring, and electrophysiological studies were obtained from medical records just before or at baseline. Rhythm abnormalities recorded by Holter or pacemaker/ICD monitoring were classified into sustained (≥ 30 s) and non-sustained (< 30 s) atrial or ventricular arrhythmias, and conduction abnormalities with sinus node dysfunction and complete heart block.

**Table 1.** , Major clinical event score developed by Woudstra et al^10^. A risk score between 0-2 corresponded to the low-risk group with a predicted 5-year risk <5%, a score between 2.5-3.5 to the intermediate-risk group with a predicted risk of 5-20%, and a score between 4-7.5 to the high-risk group with a predicted risk >20%.

| Criteria | Score<br>points |
| --- | --- |
| Age > 30 years | 1 4 |
| Repair at > 1 year | 1,5 |
| Prior ventricular arrhythmia | 1 5 |
| ≥ Moderate RV dysfunction | 1 |
| Severe tricuspid regurgitation | 1,5 6 |
| ≥ Mild LV dysfunction | 1,5 |

Baseline was determined as the first visit at the adult congenital heart disease (ACHD) center during the study period including clinical examination and 12-lead electrocardiography. Baseline data included clinical data, brain natriuretic peptide, 12-lead electrocardiography, imaging (echocardiography, cardiovascular magnetic resonance) and exercise testing performed during routine follow-up. Additional predictor variables were recorded at the time of, or just prior to baseline evaluation: specifically, unexplained syncope and episode of sustained (≥ 30 s) atrial arrhythmia. Echocardiography was considered if it was performed within the year before or after the baseline visit by an experienced operator. Echocardiographic sRV function was visually graded by cardiologists at each participating center as normal or mildly, moderately, or severely impaired ^28^. Tricuspid regurgitation (TR) severity was graded, according to the European guidelines, from absent/trivial to severe TR^28 29^. Left ventricle outflow tract obstruction (LVOT) was considered at least moderate when maximum LVOT gradient > 36mmHg or Doppler maximum velocity > 3m/s. Subpulmonary LV function was assessed from several parameters (visually estimated LV ejection fraction, fractional area change, and MAPSE) and divided into 4 groups (normal, mildly, moderately, or severely impaired) based on at least 2 parameters^19^. Brain natriuretic peptide (BNP) and cardiopulmonary exercise testing (CPET) were considered when carried out within the 3 years preceding or following the date of inclusion. Data were collected independently at participating centres and data integrity was guaranteed by each participating author.

### Study outcome

Primary endpoints were major clinical events defined by Woudstra et al. They included (1) HF events, defined as hospitalizations for HF, heart transplantation, ventricular assist device implantation, or HF as cause of death; (2) VAs, defined as symptomatic non-sustained VA, sustained VA, and sudden death; and (3) all-cause mortality.

All events occurring after patient inclusion were managed as time-dependent variables. The latest follow-up data was obtained by reviewing clinical medical records or by telephone contact and/or consultation with the patient.

### Statistical methods

Variables are described as mean ± SD, median [interquartile range (IQR)], and numbers or percentages as appropriate. Group comparisons were performed using the Student’s t-test, Mann–Whitney, or χ2 test where applicable. Follow-up time was calculated from the time of baseline evaluation to the date of reaching the study endpoint. The Kaplan–Meier method was used to estimate the incidence of reaching the study endpoint.

*Missing data*: predictive variables used in the risk score model were all available. Other data were no more missing than 1%, except for BNP, peak VO2, and predicted VO2, that were not evaluated as predictors.

*Validation of risk prediction score of major clinical events:*

Follow-up was censored at 5 years and the estimated 5-year risk of major events was calculated for each individual patient using the following equation

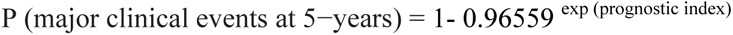

where prognostic index = score × 0.9841.

The C-index (Harrell’s) was used to measure how well the model discriminated between high-intermediate- and low-risk patients (a value of 1 indicates perfect discrimination, while a value of 0.5 indicates no discrimination)^30, 31^. Bootstrapping was used to calculate the confidence intervals (CIs). The calibration slope was used to assess the degree of agreement between the observed and predicted risk of major clinical events (a value close to 1 suggests good overall agreement)^32^. Model calibration was described graphically by stratifying patients into the three risk groups identified in Woudstra et al’study^10^.

Association of predictors with clinical events was further assessed using Cox proportional hazards method. Proportionality of hazards was confirmed in each case by assessing the correlation between the scaled Schoenfeld residuals and time. All predictors with a univariate value <0.05 were included into a multivariate model, after which stepwise backward selection based on Akaike Information Criterion (AIC) allowed to determine the best-fit model. A separate analysis was done in patients with a reduced anatomic LV function. P < 0.05 was considered statistically significant. Analyses were performed using SAS statistical software (Version 9.4, Cary, NC) and MedCalc (MedCalc Software, Mariakerke, Belgium).

### Ethics

This study complies with the Declaration of Helsinki. Local ethical approval was given for each collaborating center with waiver of informed consent for retrospective, anonymized data (NCT03833843).

## Results

### Baseline clinical characteristics

Among 512 adult patients with AtrS, 417 fulfilled inclusion criteria and constituted our study population (figure 1). Baseline characteristics are shown in Table 2. Compared to the Dutch cohort, the European cohort was younger at baseline evaluation (median age 24 years [interquartile range 18-30] vs 28 [IQR, 24–36], p<0.01), patients were more frequently operated before 1 year of age (72% vs 47%, p<0.01), and sRV function was more commonly impaired (49% vs 23%, p<0.01). Differences in baseline characteristics are summarized in Table 2.

**Figure 1.**
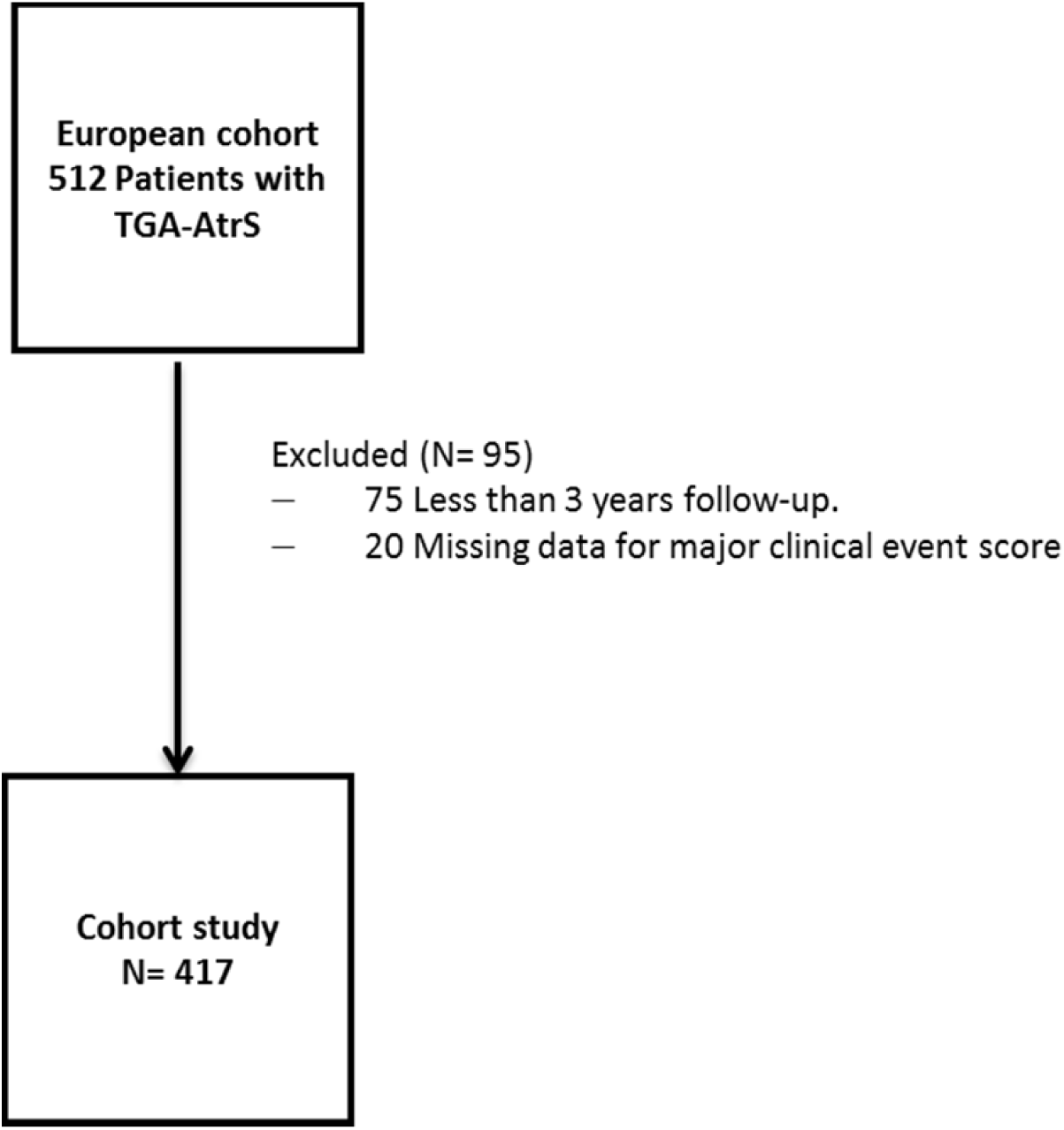
Flowchart study.

**Table 2.** Baseline Characteristics and comparison with Dutch cohort. ACEi, Angiotensin-converting enzyme inhibitor; ARA 2, Angiotensin II receptor antagonist; ICD, implantable cardioverter-defibrillator; LV, left ventricle; NYHA, New-York Heart Association; TGA, Transposition of the Great Arteries.

| **Characteristics** | **Validation cohort**<br>**(N= 417)** | **Dutch cohort**<br>**(N= 167)** | **P Value** |
| --- | --- | --- | --- |
| **Age** | 24 (18-29) | 28 (24–36) | **<0.01** |
| **Aged>30 y** | 102 (24%) | 74 (44%) | **<0.01** |
| **Gender, male** | 261 (63%) | 104 (62%) | 0.86 |
| **Senning procedure** | 212 (51%) | 66 (40%) | 0.12 |
| **Complex TGA** | 112 (27%) | 51 (31%) | 0.60 |
| **Repair > 1 year** | 115 (28%) | 89 (53%) | **<0.01** |
| **Subsequent interventions after**<br>**atrial switch surgery** | 108 (26%) | 32 (19%) | 0.41 |
| **Supraventricular tachycardia** | 140 (36%) | 58 (35%) | 0.88 |
| **Complete pregnancy** | 73 (18%) | NA | NA |
| **Pulmonary arterial hypertension** | 26 (6%) | NA | NA |
| **Prior heart failure** | 18 (4%) | 8 (5%) | 0.83 |
| **Prior ventricular tachycardia** | 38 (9%) | 13 (8%) | 0.91 |
| **NYHA I** | 304 (73%) | 128 (77%) | 0.39 |
| **NYHA ≥ II** | 112 (27%) | 39 (25%) | 0.80 |
| **Moderate or severe impairment**<br>**of RV function** | 205 (49%) | 38 (23%) | **<0.01** |
| **Severe tricuspid regurgitation** | 22 (5%) | 12 (8%) | 0.73 |
| **Mild or greater impairment of**<br>**LV function** | 35 (8%) | 8 (5%) | 0.77 |
| **Severe mitral regurgitation** | 1 (0,2%) | NA | NA |
| **Moderate pulmonary stenosis** | 55 (13%) | NA | NA |
| **Cardiac treatment** | 155 (37%) | NA | NA |
| **Beta-blockers** | 74 (18%) | 28 (17%) | 0.91 |
| **Diuretics** | 31 (7%) | 14 (8%) | 0.91 |
| **ACEi/ARA 2** | 107 (26%) | 45 (27%) | 0.90 |
| **Pacemaker** | 93 (22%) | 41 (25%) | 0.70 |
| **ICD for primary prevention** | 29 (7%) | 5 (3%) | 0.74 |

### Clinical outcomes

The validation study cohort had a median total follow-up of 11 years (IQR 8–16; range 3–19 years). Seventy-three patients reached the primary endpoint (17.5%) during the study period: 53 patients (13%) developed HF, 10 underwent heart transplantation (2%), 21 experienced ventricular arrhythmia (5%) and 15 died (4%). No patients were implanted with a ventricular assist device as destination therapy. Causes of death were HF (7), sudden cardiac death (4), non-cardiac death (2) and unknown (2). Median event-free survival of event-naive patients surviving into adulthood was 50 years (95% CI 48-52), and median survival until death or heart transplantation of adults with TGA after atrial switch was 53years (95% CI 51-54). The annual and 5-year cumulative incidence rates of major clinical events was 96.8% (95%CI 97.6-99.8) and 94.0% (95%CI 91.7-96.3) respectively (Figure 2A).

**Figure 2A.**
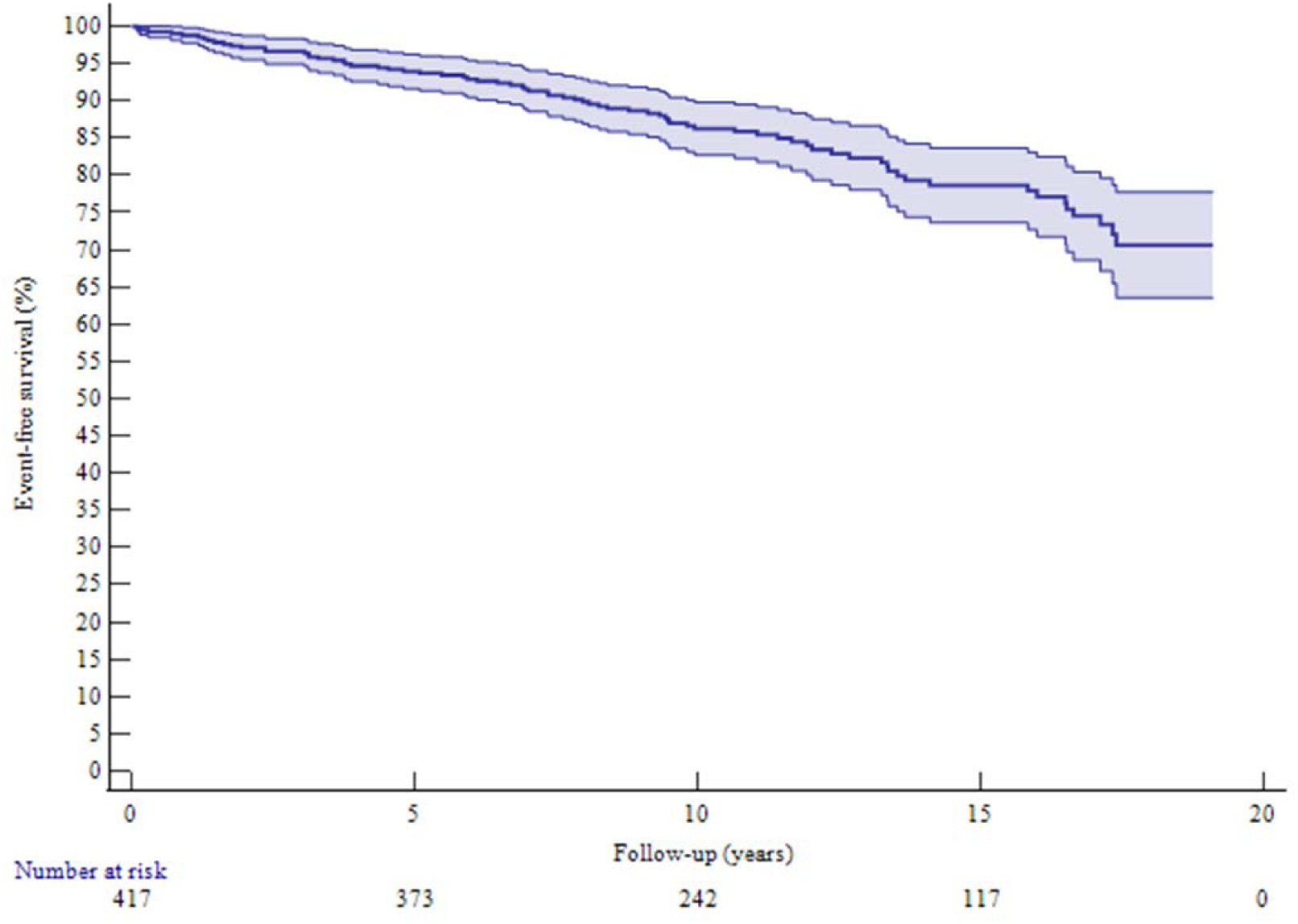
Kaplan–Meier curve showing event-free survival of the whole external validation population. Shaded area corresponds to 95% confidence interval.

According to the risk prediction model (Table 1), 298 patients had a low (71%), 92 an intermediate (22%) and 27 high (6%) risk of a major clinical events. Figure 2B shows the Kaplan-Meier event-free survival plotted and compared between low, intermediate, and high-risk group. Freedom from major clinical event survival was significantly reduced in the high- and intermediate-risk group compared to the low-risk group (log rank test, p<0.001). A significant 95% CI overlapping was observed between the intermediate and high-risk group survivals and risk of major clinical events was not significantly different between these 2 groups in the external validation population (Figure 2B, Figure 3).

**Figure 2B.**
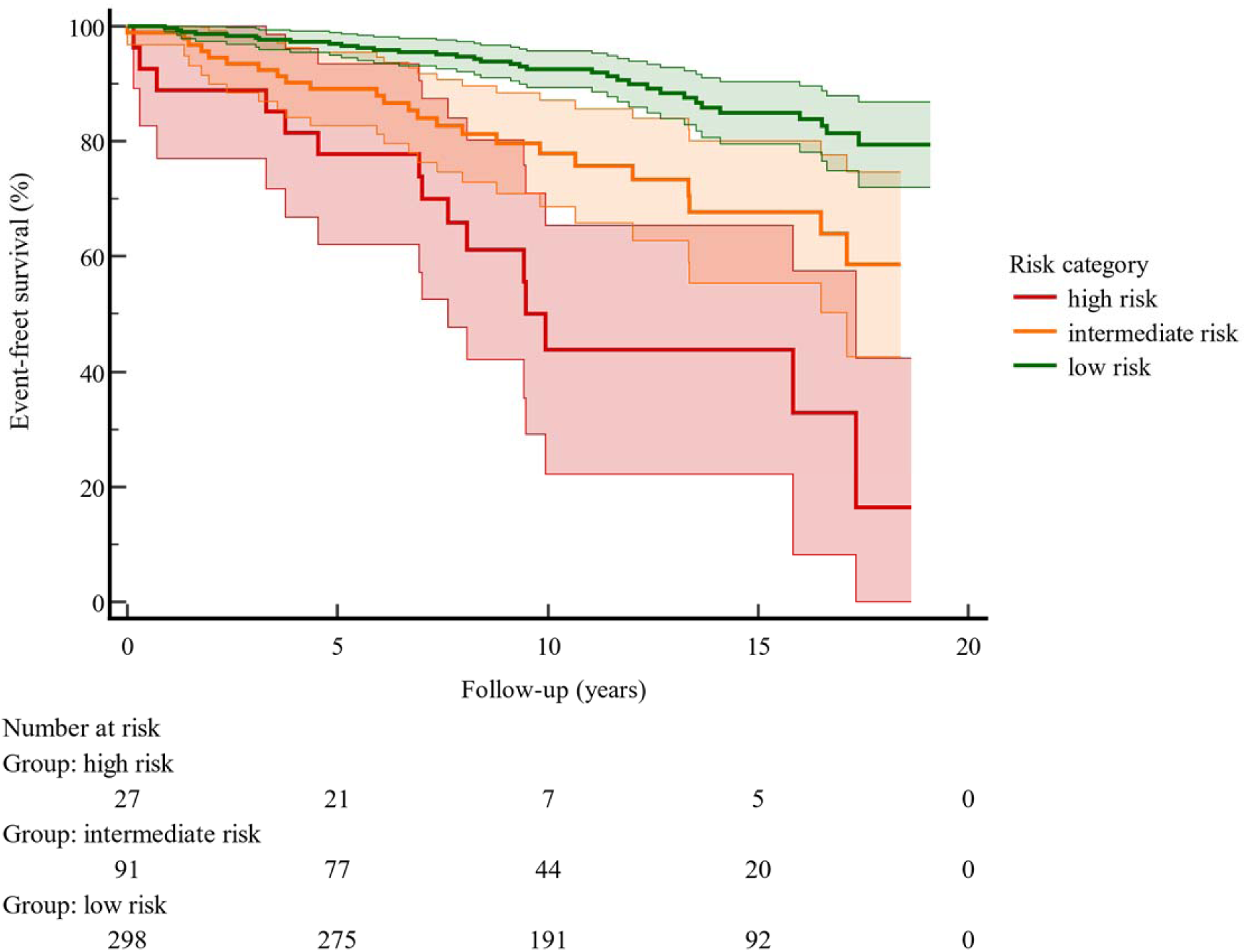
Kaplan–Meier curve showing event-free survival of the external validation cohort by risk category. Shaded areas correspond to 95% confidence intervals.

**Figure 3.**
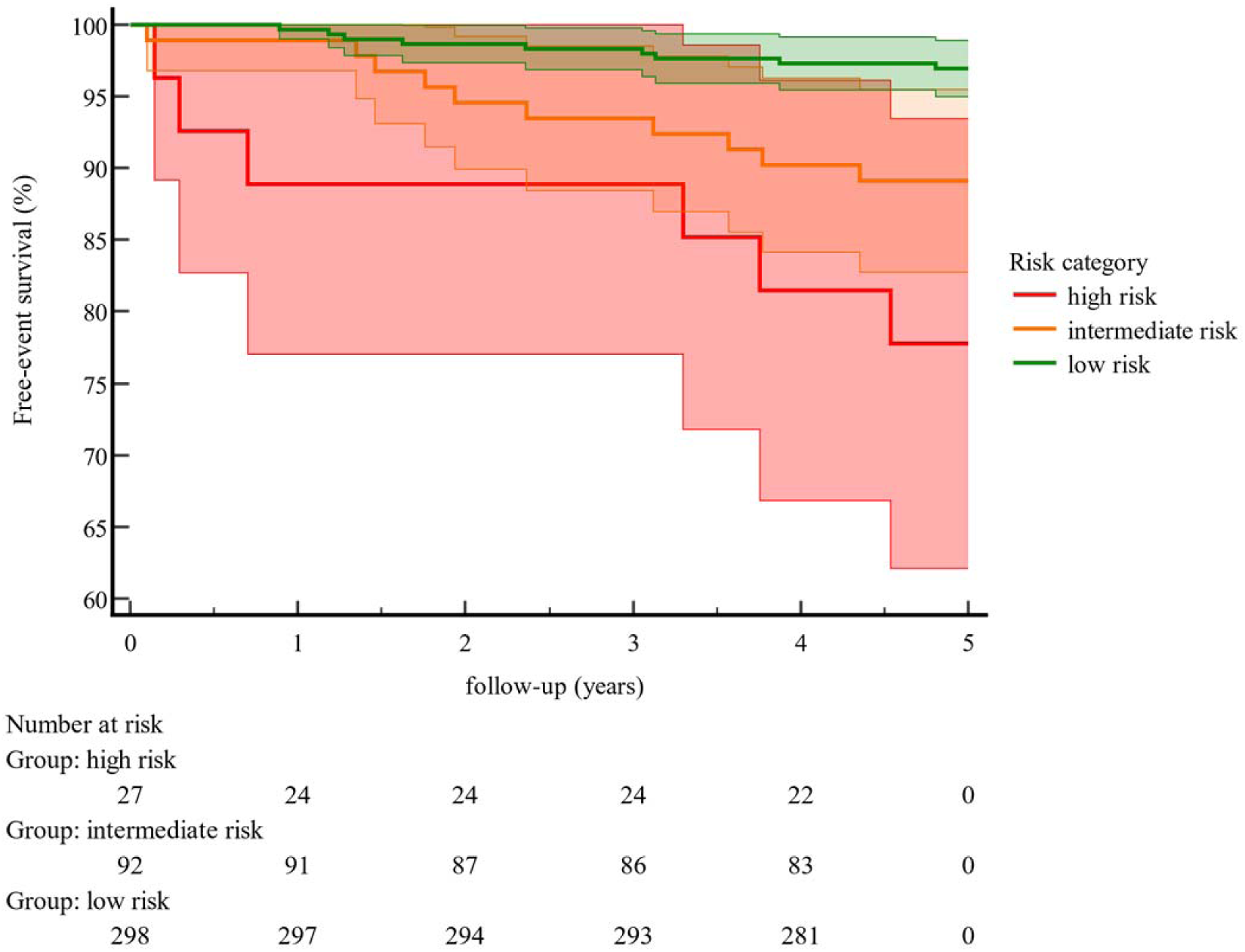
Kaplan–Meier curves showing event-free survival by risk category at 5 years. Shaded areas correspond to 95% confidence intervals.

Analyses were focused on the 25 patients who had major clinical events within 5 years of follow-up (Figure 4). Baseline characteristics in those with and without a clinical event are shown in (Table 3).

**Figure 4.**
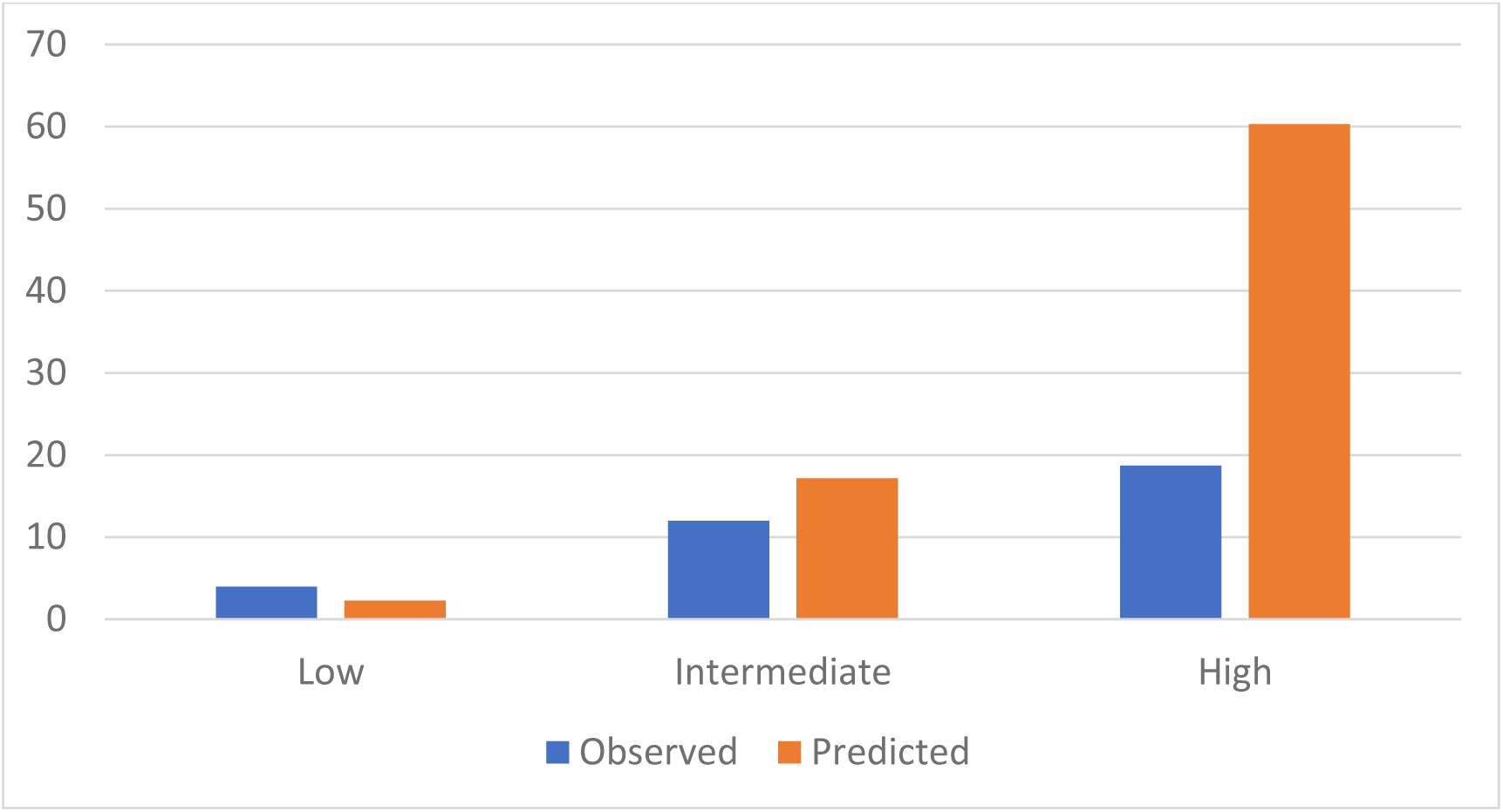
Comparison of observed and predicted risk by risk group of the major clinical events proposed risk model. Vertical bars represent model-based predicted (orange) and observed (blue) probability of events at 5 years. The low-risk group corresponds to a predicted 5-year risk <5%, the intermediate-risk group to a predicted risk of 5 - 20%, and the high-risk group to a predicted risk >20%.

**Table 3.** Baseline characteristics of patients with and without major clinical events. AtrS, atrial switch; HF, Heart Failure; LV, left ventricle; NYHA, New-York Heart Association; PAH, Pulmonary Arterial Hypertension.

| Clinical characteristic | Composite end-point (*n* = 73) | No composite endpoint (*n* = 344) | Hazard ratio (95% CI) | *P*-value |
| --- | --- | --- | --- | --- |
| Age | 27 (20-32) | 23 (18-29) | 1.09 (1.05 - 1.12) | **<0.01** |
| Gender (male) | 46 (63%) | 215 (62%) | 1.00 (0.62 - 1.61) | 0.96 |
| Aged >30y | 25 (34%) | 77 (22%) | 2.58 (1.57 – 4.23) | **<0.01** |
| Aged >1 y at AtrS | 33 (45%) | 82 (24%) | 2.41 (1.52 - 3.82) | **<0.01** |
| Mustard procedure | 44 (60%) | 161 (46%) | 1.22 (0.86 – 1.96) | 0.40 |
| Complex anatomy | 32 (44%) | 80 (23%) | 0.92 (0.49 – 1.72) | 0.81 |
| Symptoms (NYHA>1) | 30 (41%) | 83 (24%) | 2.71 (1.68 – 4.34) | **<0.01** |
| Prior HF | 56 (77%) | 16 (5%) | 30.51 (17.40 – 53.30) | **<0.01** |
| Prior VA | 12 (16%) | 26 (8%) | 1.76 0.94 - 3.26) | 0.07 |
| Prior atrial arrhythmia | 46 (63%) | 94 (27%) | 3.52 (2.18 – 5.66) | **<0.01** |
| PAH | 13 (18%) | 13 (4%) | 3.43 (1.88 – 6.26) | **<0.01** |
| Pacemaker | 31 (42%) | 62 (18%) | 2.94 (1.84 – 4.68) | **<0.01** |
| Moderate or greater RV dysfunction | 53 (73%) | 198 (56%) | 1.76 (1.05 – 2.95) | **<0.03** |
| Severe tricuspid regurgitation | 11 (15%) | 11 (3%) | 3.47 (1.83 – 6.61) | **<0.01** |
| LVOT obstruction | 16 (22%) | 39 (11%) | 1.71 (0.98 – 2.97) | 0.06 |
| Mild or greater LV dysfunction | 18 (25%) | 17 (5%) | 4.88 (2.85 – 8.33) | **<0.001** |
| Moderate or severe mitral regurgitation | 3 (4) | 2 (0.6%) | 4.78 (1.15 – 19.86) | 0.03 |
| BNP (pg/mL) | 371 (223-1007) | 130 (68-236) | 1.00 (1.00 - 1.00) | **<0.01** |
| Peak VO2 (ml/mn/kg) | 19.2 (15.7-22.5) | 23.6 (17.5-27.5) | 0.92 (0.88 – 0.97) | **<0.01** |
| VO2 predicted (%) | 47.0 (35.0-53.0) | 61.0 47.7-68.3) | 0.97 (0.94 – 0.99) | **0.01** |

### Major clinical event score model validation

The performance of the major clinical event score model in predicting risk at 5 years was assessed in 417 patients with 25 events. Harrell’s C-index was 0.73 (95% CI 0.65–0.81). The calibration slope was 0.20 (95% CI 0.03–0.36). Figure 4 illustrates the agreement between predicted and observed 5-year cumulative proportion of major clinical event for each group of predicted risk.

### Predictors of major clinical events

We separately performed an evaluation of predictors of major clinical events during study follow-up. Predictors independently associated with major clinical events are shown in Table 3. Those with a significant (p < 0.05) univariate linear relationship with the primary outcome were fitted into a multivariate model. After stepwise backward selection based on AIC, the 7 factors included in the model were age at baseline, repair after 1 year age, history of atrial arrhythmia, history of HF, NYHA >1, at least moderately sRV dysfunction and at least mild subpulmonary LV dysfunction. History of HF (HR=33.19; 95%CI 9.7-113.5; p<0.01) and at least mildly impaired sub pulmonary LV function (HR=2.96; 95%CI 1.31-6.70; p<0.01) remained the strongest predictors of major clinical events in patients with D-TGA and atrial switch.

Patients with subpulmonary LV dysfunction had more severe sRV dysfunction compared to patients without LV dysfunction (OR= 8,45; 95%CI 3,38-20,63; p<0,01). Moreover, symptoms, elevated BNP, and pulmonary hypertension were also significantly more common in patients with subpulmonary LV dysfunction (p<0.01) (Table 4).

**Table 4.** Comparison in univariate analysis of patients with and without subpulmonary left ventricular dysfunction. BNP, Brain Natriuretic Peptide; LV, left ventricle; NYHA, New-York Heart Association; PAH, Pulmonary Arterial Hypertension; sRV: systemic Right Ventricle.

| Variable | Patient with Mild or greater impairment of LV function (N = 35) | Patient without impairment of LV function (N= 382) | Odds Ratio (IC 95%) | P Value |
| --- | --- | --- | --- | --- |
| Normal RV function | 0 | 46 (12%) | 0,21 (0,01-1,32) | 0,210 |
| Mild impairment of sRV function | 12 (34%) | 154 (40%) | 0,77 (0,34-1,67) | 0,589 |
| Moderate impairment of sRV function | 11 (31%) | 160 (42%) | 0,64 (0,27-1,40) | 0,282 |
| Severe impairment of sRV function | 12 (34%) | 22 (6%) | 8,45 (3,38-20,63) | **<0,01** |
| PAH | 10 (29%) | 16 (4%) | 9,05 (3,32-23,90) | **<0,01** |
| NYHA 2-4 | 18 (51%) | 94 (25%) | 3,23 (1,50-6,98) | **<0,01** |
| BNP (N=265) | 1584 (659,25-7278) | 139,8 (78-259) | 1,00 (1,00-1,00) | **0,012** |

## Discussion

This is the first study that reports on the external validation of a major clinical event risk score in patients followed after atrial switch operation from a large, multicenter, independent European cohort. This score, developed by Woudstra et al in 2021^10^, allowed to stratify the risk of composite outcomes including HF, ventricular tachycardia, sudden death, and death. This information on individual absolute risk could help determine follow-up intensity, and support management decisions on prevention and treatment of the prevailing complications. From our externally validated cohort, we showed that this score discriminated well between high/intermediate and low-risk patients at 5 years but tended to overestimate the absolute risk of major events. Such as in Woudstra et al’ study ^10^, our results underscored the strong prognostic value of subpulmonary LV function in patients with a sRV.

Fewer major clinical events were observed in our cohort compared with Dutch study^10^. This may be explained by significant differences between our two populations. In our cohort, most patients had undergone surgery before the age of 1 year and patients were younger at study entry, which could explain our lower major clinical event rate. Even though the incidence of HF did not differ between the 2 populations (14% in the European registry vs 18% in the Dutch study), moderate to severe sRV dysfunction was more prevalent at baseline in the external validation population. This results should be cautiously interpreted because sRV function was qualitatively assessed by echocardiography, which is known to have a poor interobserver reproducibility^33 28^. However, all echocardiography exams were performed by experienced cardiologists specialized in ACHD, and the multicenter aspect of our study may have averaged the variability in the assessment of sRV function. Moreover, sRV dysfunction rate was similar to previously reported in a large cohort of 1 168 patients with AtrS operation for D-TGA^11^. It seems that unlike LV dysfunction in acquired heart disease, sRV dysfunction is far from sufficient to predict the unfavorable evolution of patient with D-TGA and atrial switch. A large number of patients have sRV dysfunction for several years without an episode of HF ^34^.

We found that the prognostic factors most associated with the occurrence of major clinical events are history of HF and subpulmonary LV dysfunction. Recently, several studies have demonstrated that adverse subpulmonary LV remodeling and systolic dysfunction were associated with worse clinical outcome in patients with a sRV ^19,22^. Subpulmonary left ventricular dysfunction is relatively common in patients with atrial switch and severe systemic right ventricular dysfunction; in our cohort, one third of patients with severe systemic right ventricular dysfunction have subpulmonary left ventricular dysfunction. Our results suggest that LV dysfunction is a sensitive sign of failing sRV circulation and underscore the importance of its evaluation in the routine assessment of patients with a sRV ^19^.

The most obvious etiologies for subpulmonary LV dysfunction appear to be RV/LV interdependence and pulmonary arterial hypertension (Table 4). Precapillary pulmonary hypertension appears to be a cause of subpulmonary LV dysfunction and is a recognized complication after atrial switch, with an estimated prevalence of 6-7% ^35^. Postcapillary pulmonary hypertension is related to atrial stiffness and dysfunction secondary to the atrial switch, tricuspid regurgitation and sRV failure^36^. Particular attention should be paid to the subpulmonary LV after atrial switch.

Our findings are complementary of those from Woudstra et al ’study^10^. Indeed, the risk model allows to identify patients with high risk of major adverse events, who will require a close monitoring in tertiary centers proposing therapeutic options for advanced HF like mechanical circulatory support and heart transplantation. This approach should mainly concern patients with intermediate or high-risk score, and even more when history of HF or at least mildly impaired subpulmonary LV function are reported. The use of prognostic scores seems essential to stratify the risk of events, particularly HF. Indeed, depending on the risk of HF, these patients may benefit from regular monitoring, cardiac rehabilitation programs, early detection and treatment of supraventricular rhythm disorders, and early HF treatment when the patient starts to be symptomatic^27^. Although medical therapy has failed to show preventive efficacy on heart failure events and death in several studies, screening of high risk patients will, maybe, allow to show some benefit in this cohort in future studies^7,8^. However, the present score models evaluate a composite endpoint, and predictors may vary according to the event assessed, even if several factors overlap the risk of HF and sudden cardiac death in patients with a sRV. Recently, a sudden death prediction score was developed for sRV (D-TGA after atrial switch and congenitally corrected transposition), based on different criteria: age, HF, syncope, severe right ventricular dysfunction, moderate pulmonary stenosis and QRS width^4^. Currently, there is no specific prognostic score for HF in this population, although it is the main cause of death in adults with D-TGA and AtrS^12^.

The main limitation of our study is the retrospective design, supported by the low incidence of events in a rare cardiac condition. By conducting a large multicentric study, bias inherent to retrospective design could be easier managed. No predictors included in the risk predictive model were missing in the external validation dataset. However, some factors known to be strong prognostic markers for clinical outcomes in sRV, such as BNP or peak VO2 measurements, were missing for more than 25% of cases and were not included in multivariable analysis. Further risk prediction models should be developed by including these markers. sRV function was not retained in multivariate analysis; however cardiac magnetic resonance imaging measurements of sRV function might show a stronger relationship with worse outcomes.

We reported the first external validation of a major clinical events risk model in a large D-TGA patient population. Although a good discrimination, the model tends to overestimate the 5-year risk of major clinical events. HF remains associated with poor outcome in this cardiac defect. Subpulmonary LV dysfunction appears to be a key marker in the prognosis of patients with Senning and Mustard. The development or optimizing new risk models is required, particularly to predict HF and individualize the management of subgroups of patients with D-TGA after AtrS.

## Data Availability

The data will not be made available to other researchers for purposes of reproducing the results or replicating the procedure

## Non-standard Abbreviations and Acronyms

D-TGA: Transposition of the great arteries
AtrS: Atrial switch
sRV: Systemic right ventricular
CHD: Congenital heart disease
LVOT: Left ventricle outflow tract
VSD: Ventricular septal defect

## Acknowledgements

We thank Mrs Anissa Boubrit, a French research technician from the reference center of complex congenital heart disease M3C, who helped with recording of data from the Paris.

## Supplemental Material

Table S1

